# Prevalence and predictors of indoor second-hand smoke among the non-smoker household population in India: Evidence from the Demographic and Health Survey (DHS) 2021 data

**DOI:** 10.64898/2026.01.18.26344347

**Authors:** Vishal R. Tikhute, Komal D. Devkate, Prashant Gawai

## Abstract

Despite high risk, exposure to indoor second-hand smoke (SHS) among the non-smokers is scarcely studied in India. To address this gap, we assessed the prevalence and predictors of indoor SHS among the non-smoker household population of India. Bivariate and multivariate analysis of Demographic and Health Survey 2021 data of 1,529,486 household members was carried out. For India, the prevalence of indoor SHS exposure was 40.6 percent. Adults (aOR 0.88; 95% CI 0.87–0.89), educated individuals (aOR 0.87; 95% CI 0.86–0.88), married couples (aOR 0.72; 95% CI 0.71–0.73), and occupants of pucca houses (aOR 0.77; 95% CI 0.75–0.78) had lower odds of indoor SHS exposure than their respective reference categories. Females (aOR 1.54; 95% CI 1.53–1.56), higher household size (aOR 1.07; 95% CI 1.07–1.08), joint family (aOR 1.11; 95% CI 1.10–1.12), Muslims (aOR 1.18; 95% CI 1.17–1.20), Scheduled Tribes (aOR 1.15; 95% CI 1.14–1.17), rural residents (aOR 1.26; 95% CI 1.25–1.27), and the northeastern zone (aOR 1.12; 95% CI 1.10–1.14) were positively associated with indoor SHS exposure. Every four out of ten Indians experienced indoor SHS exposure. Half of them were daily exposed to indoor SHS. Particularly children, women, and occupants of *Kachha* houses were at higher risk. The susceptibility among these high-risk groups to cancers, respiratory diseases, and cardiovascular diseases can be averted by improving housing, increasing access to tobacco cessation services and additional pharmacotherapies, and strict enforcement of tobacco control regulations in India.

## Introduction

Second-hand smoke (SHS) is a combination of smoke given off by the burning end of a tobacco product and the smoke exhaled by the smoker (1,2). SHS contains more than 7,000 pollutants, and about 40 of them are carcinogens (1–3). SHS exposure is closely related to multiple health issues such as lung cancer, cardiovascular diseases, adverse reproductive health effects in women (e.g., low birth weight), metabolic diseases (Type II Diabetes Mellitus), and other indoor air pollution-related - health issues (3–7). SHS exposure can also lead to premature deaths among non-smokers (3). In 2019, SHS exposure caused 1.3 million deaths worldwide (8,9). Most of the deaths were reported in low- and middle-income countries (LMIC) (8,9). The burden and correlates of SHS among children, adolescents, and adults have been widely discussed in the context of selected African (10–12), European (13), American (14–18), and some Asian countries (19– 22). Most of these studies covered school children and SHS among adults in the vehicles or at work or public places (14–17,21,22). None of them covered the recent situation of indoor SHS in India.

After China, India is the second largest producer and consumer of tobacco products (23,24). According to the Global Adult Tobacco Survey (GATS 2), 28.6% of 15 and above Indian population use tobacco in any form (25). Tobacco use, contributes to annually 1.35 million preventable deaths in India (24,26,27). As the Cigarettes and Other Tobacco Products (Prohibition of Advertisement and Regulation of Trade and Commerce, Production, Supply and Distribution) Act, or COTPA 2003 restricts smoking in public places (28,29). However, there are no restrictions on smoking inside homes. This results in displacement effect of smoke-free legislation evidenced in China (30), where smokers prefer smoking inside their households as smoking is banned outside. Resultantly, due to indoor SHS, smoking-related risk is distributed equally among household members, including non-smoker adults and children.

Although smokeless tobacco is preferred in India (23), comparatively, smoking is more hazardous than smokeless tobacco (31,32). However, despite higher susceptibility, only a few studies attempted assessing the magnitude of SHS at public places (33,34). Further, some small-scale studies covering specific cities assessed the indoor SHS exposure among children (35,36), urban youth (18-21 years) (37,38), particularly in public places (39), None of these studies has assessed the national-level burden of indoor SHS exposure among household members of all age groups by analyzing the most recent India Demographic and Health Survey (DHS) 2021 data. Further, the earlier literature has not assessed the sociodemographic factors associated with indoor SHS exposure among non-smokers. Therefore, considering the susceptibility of the non-smoker household population of all ages to health risks associated with indoor SHS, it is crucial to assess the prevalence and socio-demographic predictors of indoor SHS exposure in India. The present study aims to assess the prevalence and predictors of indoor SHS exposure among the household population in India using recent DHS 2021 data.

## Materials and Methods

A cross-sectional study design was used to assess the prevalence and predictors of indoor SHS exposure in India using DHS 2021 data (also known as the national family health survey, NFHS-5) (40).

The authors of this study have obtained the authorization letter from the DHS program, allowing use of IRB approved India DHS 2021/NFHS-5 data for this study. The DHS authorization exempts obtaining separate IRB approval for this study, as it doesn’t involve any clinical data with individual identifiers. The data used in this study is anonymous, collective population data that doesn’t provide information on individual identifiers. The study has followed all terms and conditions of data use specified on the DHS program’s website (41).

### Data and sampling

This study used data from the household member recode dataset (IAPR7EDT.DTA file) of NFHS-5 (40). This is the most recent round of the NFHS, which covered a total population of 2,843,917 household members from all six regions and 36 states and UTs of India. The NFHS-5 provides information on population, health, and nutrition for India, covering all 36 states and union territories (UT) and 707 districts (42). The peculiarity of NFHS-5 is that it also provides data on the use of substances like tobacco and alcohol.

The study used two-stage purposive sampling method. As indoor smoking affects each member of the household, therefore, at the first stage, a total population of 2,843,917 all age household members, including children, were selected. Further in second stage, from these 2,843,917 individuals, 1,303,843 (45.8% of the total population) entries for the smoker population were removed. Further, 10,588 (0.4% of total population) entries for missing values and ‘don’t know’ responses were removed. The rest of the entries covering a total of 1,529,486 non-smokers household members were used as a study sample (Figure 1).

**Figure 1.**
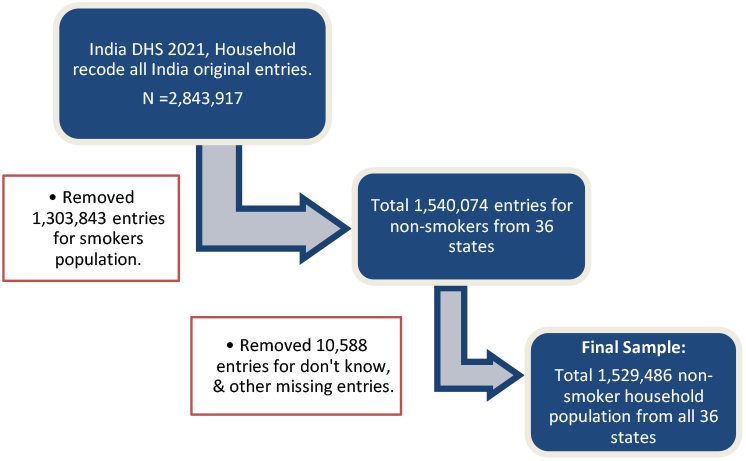
Strategy for selecting sample from India DHS 2021 data

### Outcome variable

The NFHS-5 household recode dataset had a categorical variable ‘hv252’ i.e., ‘frequency of household members smoking inside the house’ which originally had four response categories (i.e., 0 = never, 1 = daily, 2 = weekly, 3 = monthly, and 4 = less than once a month).

This variable ‘hv252’ was recoded into the new dichotomous variable ‘indoor SHS exposure’. For this, the original category ‘0’ was recoded as ‘no’ and the rest of the four categories (1 to 4) were recoded as ‘yes’ (the final responses for the new variable were ‘no’ or ‘yes’). This new variable, i.e., indoor SHS exposure (yes or no), representing the exposure to indoor SHS, was considered a dependent variable for further analysis.

### Explanatory variables

To have sufficient representation (minimum 5%) for each categorical value, three raw variables, i.e., educational attainment (four categories), religion (ten categories), and current marital status (five categories), were recoded into three new variables (with smaller categories). Raw variable ‘educational attainment’ (four categories) was recoded into dichotomous variable ‘education’ (no education and educated). Raw variable ‘current marital status’ (four categories) was recoded into dichotomous variable ‘marital status’ (never married or ever married). And another variable religion (ten categories) was recoded into a new variable ‘religion’ (Hindu, Muslim, Christian, no religion, and others).

Further, to present vulnerability among children, a new dichotomous variable age group (under 18 or 18 and above) was generated from raw continuous variable age. The age criteria of 18 years were used, as in India a child is defined as anyone under 18 years of age. Additionally, as India has six geographical regions (zones) covering all 36 states/UTs, accordingly, states are grouped into these six zones: northeast (NE), northern (N), eastern (E), central (C), western (W), and southern (S). Therefore, variable ‘states’ was recoded into variable “zone” (six categories). The list of explanatory independent variables also included five raw variables such as type of house (*kachha*, semi-pucca, and *pucca*), social category (scheduled caste, scheduled tribe, other backward class, and general), wealth index (poorest, poorer, middle, richer, and richest), place of residence (urban and rural), and alcohol use (no or yes).

### Data analysis

The data were analyzed using Python (version 3.14.2) (43) with Spyder IDE (44), employing libraries such as pandas and statsmodels for statistical analysis. Additionally, QGIS v3.14 was used to prepare a map presenting the state-specific prevalence of indoor SHS. Descriptive, bivariate, and binomial logistic regression analyses were carried out to assess the prevalence and predictors of indoor SHS exposure in India.

For bivariate analysis, a first-step chi-square test was performed to determine factors correlated with indoor SHS. For hypothesis testing, *p* < 0.05 was considered significant. Any variable significantly associated (*p*<0.05) with the outcome variable was considered for further multivariate analysis.

A binomial logistic regression was carried out to assess the factors associated with indoor SHS exposure in India. Using the adjusted odds ratios (aOR), the association between the outcome variable and explanatory variables was presented. The goodness-of-fit of the model was assessed using the Hosmer-Lemeshow test. Further, the Akaike Information Criterion (AIC) and the Bayesian Information Criterion (BIC) were calculated. The state-wise regional distribution of indoor SHS was presented using GIS mapping (45).

## Results

The mean age of the study population was 37 years (SD ±17.1), and 90 percent of the population was above 18 years (Table 1). The sample covered 61 percent males and 39 percent females. About 77 percent of individuals were educated. About 72 percent of individuals were ever married. About 54 percent of individuals lived in non-nuclear joint families. Sixty percent of individuals lived in pucca houses. About three-fourths of individuals were Hindus. About 41 percent belonged to the other backward class, followed by the general (23%), scheduled caste (20%), and scheduled tribes (16%). About 37 percent of the individuals belonged to the poorest and poorer wealth quintiles. Seventy-two percent of individuals lived in rural areas. Considering the behavioral components, the prevalence of alcohol drinking in India was four percent (95% CI 4.1-4.2).

**Table 1.** Details of the non-smoker household population in India (N= 1,529,486)

| Variables | n | % |
| --- | --- | --- |
| **Age, Mean (SD)** | 37 (SD ±17.1) | - |
| **Age groups** |  |  |
| Under 18 | 148,901 | 9.7 |
| 18 & above | 1,380,585 | 90.3 |
| **Sex** |  |  |
| Male | 592,626 | 38.8 |
| Female | 936,860 | 61.3 |
| **Education** |  |  |
| No Education | 355,451 | 23.2 |
| Educated | 1,174,035 | 76.8 |
| **Marital status** |  |  |
| Never Married | 429,662 | 28.1 |
| Ever Married<sup>a</sup> | 1,099,824 | 71.9 |
| **Household size, Mean (SD)** | 5.3 (SD ±2.5) | - |
| **Family type** |  |  |
| Nuclear | 700,291 | 45.8 |
| Non-nuclear | 829,195 | 54.2 |
| **Type of house<sup>b</sup>** |  |  |
| Kachha | 69,448 | 4.6 |
| Semi-pucca | 526,425 | 34.8 |
| Pucca | 915,882 | 60.6 |
| **Religion** |  |  |
| Hindu | 1,170,792 | 76.6 |
| Muslim | 186,160 | 12.2 |
| Christian | 88,612 | 5.8 |
| Others<sup>c</sup> | 83,922 | 5.5 |
| **Social category<sup>d</sup>** |  |  |
| Scheduled caste | 295,380 | 20.2 |
| Scheduled tribe | 233,466 | 15.9 |
| Other backward class | 599,223 | 40.9 |
| General | 337,314 | 23.0 |
| **Wealth Index** |  |  |
| Poorest | 256,483 | 16.8 |
| Poorer | 306,093 | 20.0 |
| Middle | 317,908 | 20.8 |
| Richer | 320,027 | 20.9 |
| Richest | 328,975 | 21.5 |
| **Place of residence** |  |  |
| Urban | 420,475 | 27.5 |
| Rural | 1,109,011 | 72.5 |
| **Zone** |  |  |
| Northern | 321,412 | 21.0 |
| North-Eastern | 162,164 | 10.6 |
| Central | 377,519 | 24.7 |
| Western | 163,732 | 10.7 |
| Southern | 280,194 | 18.3 |
| Eastern | 224,465 | 14.7 |
| **Indoor SHS** |  |  |
| No | 908,825 | 59.4 |
| Yes | 620,661 | 40.6 |
| **Frequency of indoor smoke** |  |  |
| Daily | 337,941 | 54.5 |
| Weekly | 134,791 | 21.7 |
| Monthly | 79,167 | 12.8 |
| Less than once a month | 68,762 | 11.1 |
| **Alcohol use** |  |  |
| No | 1,465,090 | 95.8 |
| Yes | 64,396 | 4.2 |

<sup>a</sup> Includes ever married individuals (widowed, separated, and divorced). <sup>b</sup> n = 1,511,755; rest of the missing 17,731 entries were excluded from the analysis.
<sup>c</sup> Others = Jains, Sikhs, Jews, Buddhists, and Zoroastrians. <sup>d</sup> n = 1,465,383; missing 64103 entries were excluded from the analysis.

Among individuals whose household members smoked inside the house (*n* = 620,661), for 54.5 percent of such individuals, household members smoked daily inside their house. This was followed by individuals whose household members smoked once a week (22%), once a month (13%), and less than once a month (10.5%) inside their house.

North-eastern (58%), central (50%), and northern (48%) zones each had prevalence higher than the national prevalence. The prevalence for eastern and western zones was 36 percent and 31 percent, respectively. The southern zone had reported the lowest prevalence among all six Indian zones (18%) (Table 2).

**Table 2.** Zone-wise distribution of indoor SHS in India.

| State/Zone | n | % | Population |
| --- | --- | --- | --- |
| Jammu & Kashmir | 35,339 | 68.0 | 51,945 |
| Rajasthan | 57,179 | 64.5 | 88,586 |
| Ladakh | 3,278 | 62.9 | 5,213 |
| Haryana | 27,967 | 49.9 | 56,038 |
| Himachal Pradesh | 12,941 | 48.0 | 26,940 |
| NCT of Delhi<sup>a</sup> | 7,366 | 27.0 | 27,234 |
| Punjab | 10,514 | 16.7 | 63,066 |
| Chandigarh | 322 | 13.5 | 2,392 |
| **Northern (Total)** | **154,906** | **48.2** | **321,414** |
| Mizoram | 4,627 | 75.2 | 6,157 |
| Meghalaya | 12,980 | 74.3 | 17,459 |
| Arunachal Pradesh | 22,568 | 66.3 | 34,021 |
| Tripura | 6,022 | 66.2 | 9,096 |
| Manipur | 7,298 | 63.5 | 11,501 |
| Nagaland | 9,002 | 49.4 | 18,221 |
| Assam | 28,858 | 49.1 | 58,773 |
| Sikkim | 2,932 | 42.3 | 6,936 |
| **North-Eastern (Total)** | **94,287** | **58.1** | **162,164** |
| Uttar Pradesh | 98,351 | 52.2 | 188,570 |
| Uttarakhand | 16,180 | 52.1 | 31,074 |
| Chhattisgarh | 25,462 | 47.8 | 53,280 |
| Madhya Pradesh | 48,981 | 46.8 | 104,595 |
| **Central (Total)** | **188,974** | **50.1** | **377,519** |
| Gujarat | 26,188 | 35.7 | 73,385 |
| Maharashtra | 21,937 | 27.8 | 78,834 |
| DNH & DD<sup>b</sup> | 1,362 | 21.9 | 6,206 |
| Goa | 1,030 | 19.4 | 5,311 |
| **Western (Total)** | **50,517** | **30.9** | **163,736** |
| Andhra Pradesh | 6,714 | 24.1 | 27,881 |
| Telangana | 15,673 | 23.9 | 65,698 |
| Karnataka | 13,510 | 18.7 | 72,320 |
| A & N Islands<sup>c</sup> | 516 | 15.5 | 3,327 |
| Kerala | 4,341 | 13.1 | 33,016 |
| Tamil Nadu | 7,690 | 11.7 | 65,641 |
| Lakshadweep | 297 | 11.6 | 2,568 |
| Puducherry | 1,035 | 10.6 | 9,743 |
| **Southern (Total)** | **49,776** | **17.8** | **280,194** |
| West Bengal | 19,442 | 50.1 | 38,809 |
| Bihar | 32,916 | 39.0 | 84,481 |
| Jharkhand | 18,250 | 35.9 | 50,854 |
| Odisha | 11,593 | 23.0 | 50,322 |
| **Eastern (Total)** | **82,201** | **36.6** | **224,466** |
| **India (Total)** | **620,661** | **40.6** | **1,529,493** |

<sup>a</sup> NCT = National Capital Territory of Delhi. <sup>b</sup> DNH & DD = Dadra and Nagar Haveli and Daman and Diu. <sup>c</sup> A & N = Andaman & Nicobar Islands.

The prevalence of indoor SHS among all 36 Indian states and UTs ranged from the highest in Mizoram (75%) to the lowest in Puducherry (10.6%) (Figure 2). Except for alcohol use, the rest of the explanatory variables showed a significant association with the outcome variable at bivariate analysis (chi-square test) (Table 3).

**Table 3.** Bivariate analysis of factors associated with indoor SHS exposure in India.

| Variable | % | Chi square | p value |
| --- | --- | --- | --- |
| **Age group** |  | 6.3e+03 | 0.000* |
| Under 18 | 50.2 |  |  |
| 18 & above | 39.5 |  |  |
| **Sex** |  | 1.50E+04 | 0.000* |
| Male | 34.4 |  |  |
| Female | 44.5 |  |  |
| **Education** |  | 6.30E+03 | 0.000* |
| No Education | 46.3 |  |  |
| Educated | 38.8 |  |  |
| **Marital status** |  | 6.70E+03 | 0.000* |
| Never Married | 45.8 |  |  |
| Ever Married<sup>a</sup> | 38.6 |  |  |
| **Family type** |  | 3.20E+03 | 0.000* |
| Nuclear | 38.1 |  |  |
| Non-nuclear | 42.6 |  |  |
| **Type of house** |  | 3.90E+04 | 0.000* |
| Kachha | 54.4 |  |  |
| Semi-pucca | 49.6 |  |  |
| Pucca | 34.2 |  |  |
| **Religion** |  | 7.40E+03 | 0.000* |
| Hindu | 39.9 |  |  |
| Muslim | 44.9 |  |  |
| Christian | 49.5 |  |  |
| Others<sup>b</sup> | 31.6 |  |  |
| **Social category** |  | 2.20E+04 | 0.000* |
| Scheduled caste | 42.8 |  |  |
| Scheduled tribe | 53.0 |  |  |
| Other backward class | 36.6 |  |  |
| General | 36.1 |  |  |
| **Wealth Index** |  | 4.70E+04 | 0.000* |
| Poorest | 51.3 |  |  |
| Poorer | 48.7 |  |  |
| Middle | 42.1 |  |  |
| Richer | 36.2 |  |  |
| Richest | 27.5 |  |  |
| **Place of residence** |  | 2.80E+04 | 0.000* |
| Urban | 29.8 |  |  |
| Rural | 44.7 |  |  |
| **Zone** |  | 1.10E+05 | 0.000* |
| Northern | 48.2 |  |  |
| North-Eastern | 58.1 |  |  |
| Central | 50.1 |  |  |
| Western | 30.9 |  |  |
| Southern | 17.8 |  |  |
| Eastern | 36.6 |  |  |
| **Alcohol use** |  |  |  |
| No | 40.6 | 0.4664 | 0.495 |
| Yes | 40.7 |  |  |

\*Significant at $p < 0.05$ . <sup>a</sup> Includes ever married individuals (widowed, separated, and divorced). <sup>b</sup> Others = Jains, Sikhs, Jews, Buddhists, and Zoroastrians.

**Figure 2.**
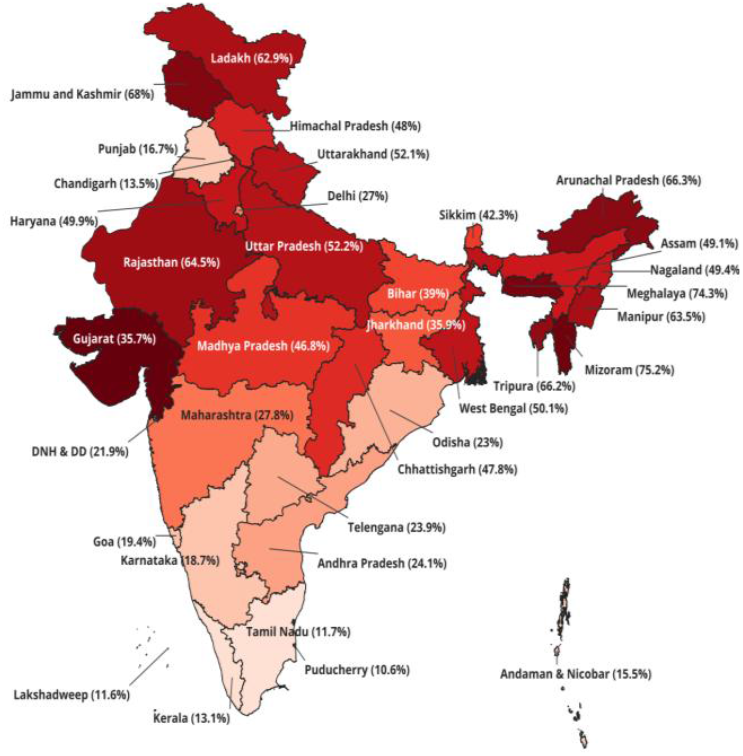
Prevalence of indoor SHS exposure among non-smoker household population in 36 Indian states

All such variables that showed a significant association (*p* < 0.05) were further entered into the multivariate (binomial) logistic regression model (Table 4). The statistical association using adjusted odds ratios shows that age group, sex, education, marital status, household size, family type, religion, social category, wealth index, residency, and zones had significant association with indoor SHS exposure in India.

**Table 4.**
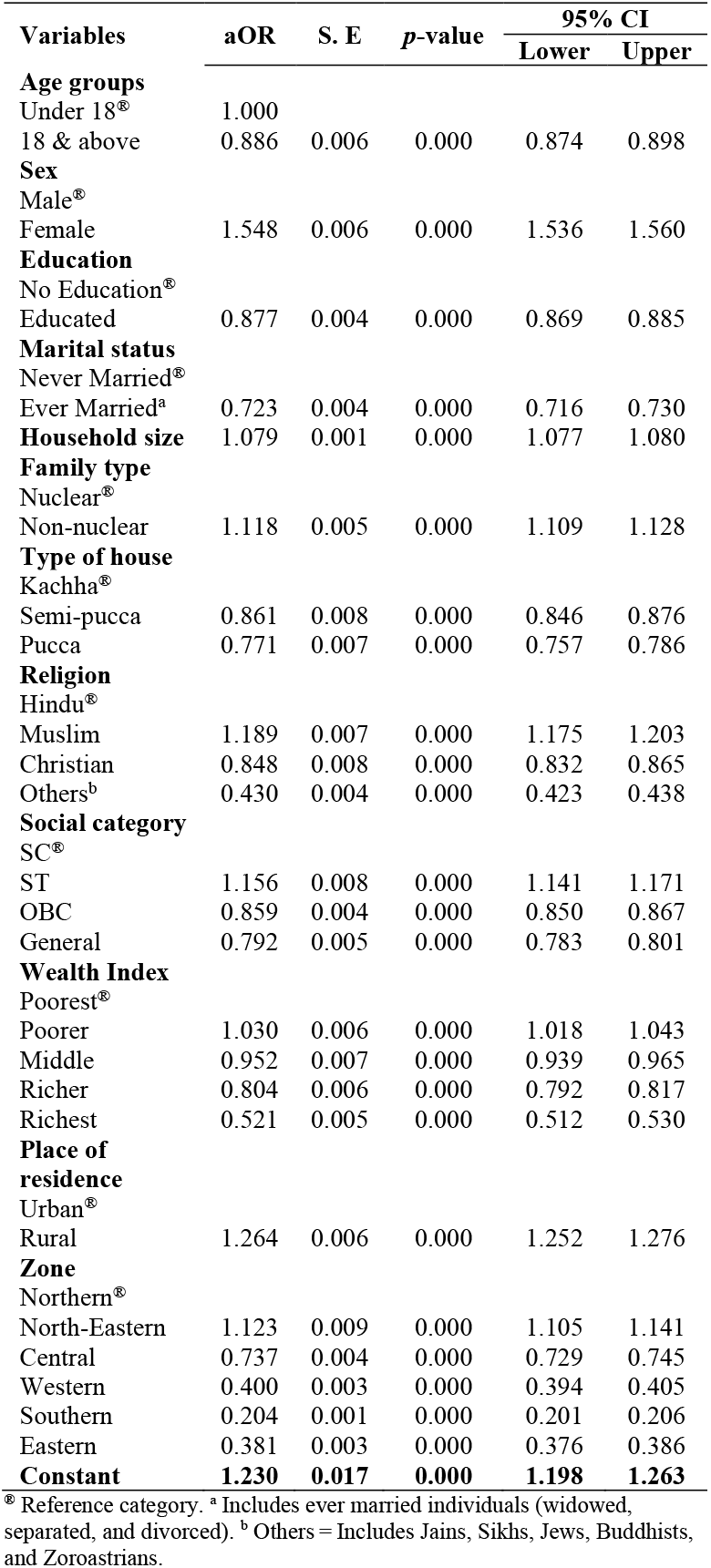
Predictors of indoor SHS among non-smoker household population in India.

Those aged 18 years and above (adults) were less likely to get indoor SHS exposure than the under-18 population (aOR 0.88; 95% CI 0.87– 0.89). This suggests that children under 18 years of age were more susceptible to indoor SHS exposure than adults. Females, compared to males, were more likely to get indoor SHS exposure (aOR 1.54; 95% CI 1.53–1.56).

Educated individuals (aOR 0.87; 95% CI 0.86– 0.88) were less likely to get indoor SHS exposure than uneducated individuals (no education). The odds of indoor SHS were lower among ever-married individuals (aOR 0.72; 95% CI 0.71–0.73) than unmarried (never married) individuals.

With the addition of one member to the household, the odds of indoor SHS exposure increased by 7 percent (aOR 1.07; 95% CI 1.07-1.08). This clearly conveys the significance of higher household size (leading to overcrowding), increasing the likelihood of indoor SHS exposure - among household members. Likewise, as compared to nuclear families, individuals from joint (non-nuclear) families were more likely to get indoor SHS exposure (aOR 1.11; 95% CI 1.10–1.12). Compared to *kachha* houses, occupants of semi-pucca (aOR 0.86; 95% CI 0.84–0.87) and pucca (aOR 0.77; 95% CI 0.75–0.78) houses were less likely to get indoor SHS exposure.

Muslims comparatively had higher odds of indoor SHS than Hindus (aOR 1.18; 95% CI 1.17– 1.20). While Christians (aOR 0.84; 95% CI 0.83– 0.86) and individuals from other minority religions (aOR 0.43; 95% CI 0.42–0.43) were less likely to get indoor SHS exposure than Hindus. STs were more likely to get indoor SHS exposure than SCs (aOR 1.15; 95% CI 1.14–1.17).

Except for the poorer wealth quintile (aOR 1.03; 95% CI 1.01–1.04), individuals from the rest of the wealth quintiles were less likely to get exposed to indoor SHS than the poorest quintiles. As compared to the urban population, individuals from rural areas (aOR 1.26; 95% CI 1.25–1.27) were more likely to get indoor SHS exposure.

As compared to the northern region, the north-eastern region (NER) had higher odds of getting indoor SHS exposure (aOR 1.12; 95% CI 1.10– 1.14). However, comparatively, the risks of indoor SHS were lower among individuals from other regions than those from the northern region.

## Discussion

The study assessed the burden of indoor SHS across all 36 Indian states and UTs. Additionally, socio-demographic factors associated with indoor SHS among the Indian household population were identified. The previous literature has covered specific sections of the population, such as adults at work or in public places; no study covered the household population of all age groups, including children, which has a higher risk of direct and prolonged exposure to SHS. Health risks such as - cardiovascular diseases, additional risks of smoking susceptibility (i.e., inclination to initiate cigarette smoking) (10), and reduced cognitive performance among never-smoking schoolchildren have been documented elsewhere (46). Additionally, never-smoker adolescents exposed to SHS endorsed nicotine dependence symptoms (47). Therefore, it is crucial to understand SHS exposure among children, their household dynamics, patterns, and frequency of indoor SHS exposure in detail. To address this gap, the current study has covered all age populations of non-smokers, including children, and found that the prevalence of indoor SHS exposure was significantly higher among under-18 (50%) than adults (39.5%). Suggesting that every second Indian household child is exposed to indoor SHS and is susceptible to cardiovascular diseases, smoking susceptibility and reduced cognitive performance.

The prevalence of indoor SHS exposure in India is higher than prevalence estimates that were presented for other South and Southeast Asian countries (46,48,49). Additionally, the prevalence of indoor SHS exposure assessed in the present study (40.6%) is similar to the prevalence estimated by Chawla (50), who also analyzed NFHS-5 data. However, the gender-specific prevalence differed; Chawla (50) reported higher values of gender-specific prevalence than the present study. Chawla (50) reported higher prevalence among males (62.1%) than females (47.7%) (50). While in the present study, females had higher (44.5%) prevalence than males (34.4%). These differences are attributed to different sampling approaches and analytical frameworks used in both studies. Chawla (50) had covered only the 15-49-year population (both smokers and non-smokers) and presented results using the descriptive statistics. While the present study assessed the burden of indoor SHS exposure specifically among non-smokers (of all ages) and also assessed factors contributing to indoor SHS exposure using multivariate analysis.

The socio-demographic factors associated with indoor SHS included age group, sex, education, marital status, number of household members, religion, caste, place of residence, type of house, type of family, and zones. Particularly, children, women, uneducated individuals, and rural residents who lived in joint (non-nuclear) families are at higher risk of indoor SHS exposure. Therefore, these groups need to be considered an ‘at-risk’ population. For instance, as children usually spend most of the time inside the house, they are at risk of getting exposed to indoor SHS. Further, smoking susceptibility (51,52) among children (i.e., smoking experimentation and initiation by imitating adults) increases if they are exposed to indoor SHS (21,22,30,52,53).

The gender difference in indoor SHS exposure found in the current study can be linked to multiple socio-cultural practices such as patriarchy-based gender roles, where women mostly work inside the house (54). Therefore, exposure to indoor pollution (including indoor SHS) is higher among women (55).

With the increase in the number of household members (leading to overcrowding), the exposure to indoor SHS increased. Particularly, rural residents, joint family members, those belonging to the poorest wealth quintile, and those living in congested *kachha* houses with higher household size were exposed to indoor SHS. This is in alignment with studies conducted in other low- and middle-income countries that pointed out a higher risk of cardiovascular and other respiratory diseases attributed to indoor SHS exposure intensified by poor housing conditions and poverty (11,34,56– 58). Considering this, there is a need to improve housing and overall living conditions, particularly in rural India.

Targeted interventions should be designed to increase awareness about the harms and prevention of indoor SHS among at-risk populations. Screening of these at-risk population groups against potential adverse health consequences, such as cancer, can help with early identification and timely management. At the clinical level, the exposure of individuals to carcinogens can be monitored by urinary and DNA adduct biomarkers (32). This can help with the early detection of individuals at high risk for cancer. Replicating this at the community level, through mass screening campaigns, can be an effective strategy in cancer prevention (32). This can be an effective step towards preventing tobacco-smoke-related health harms (including cancer) among smokers as well as non-smokers.

Additionally, at the macro and micro levels, there is a need for informed, targeted strategies. A recent welcome move is the inclusion of nicotine replacement therapy (NRT) in the National List of Essential Medicines (NLEM), making NRT accessible and affordable through health insurance coverage (59). This will help occasional smokers to quit cigarette smoking. Though the addition of NRT to the NLEM is an important milestone toward smoking cessation interventions in India, it covers only a limited population of cigarette smokers; it’s not effective in the case of Bidi users and light smokers (55). The NRT has a limited effect on daily light smokers (cigarettes per day; CPD < 10); therefore, there is a need to include additional pharmacotherapies (like bupropion, varenicline, etc.) in the NLEM (59). The NRT has a limited effect on daily light smokers (cigarettes per day; CPD<10); therefore there is need include of additional pharmacotherapies (like bupropion, varenicline, etc.) in the NLEM (59). This will help tobacco users to quit smoking at all levels. Studies showed that the combination of behavioral treatment and cessation medications has a higher impact on tobacco cessation than only the use of NRT (59–63,63–66).

At the individual and clinician levels, informed intervention strategies using the 5 As approach can be useful (67,68). Where clinicians should Ask all clinic-visiting adults about tobacco use; advise them to quit smoking through clear, personalized messages; assess their willingness to quit; assist them in quitting; and arrange follow-up and support for them (67,68). Additionally, cessation interventions utilizing mobile devices and social media have also shown promising results elsewhere (69,70). Brief advice from a healthcare worker, telephone helplines, automated text messaging, and printed self-help materials can also facilitate smoking cessation (2,63). Further, while some individuals smoke to avoid withdrawal effects and others smoke to get high, therefore cessation services should be customized to meet the individual needs (69). Additionally, there is a need for creating enabling environments such as brief advice, anti-tobacco messages, and strict enforcement of tobacco control laws in India (59,69).

India is one of the countries that acceded to the WHO Framework Convention on Tobacco Control (FCTC) in 2004 (71,72). Accordingly, smoking is prohibited in selected public places (but not at homes). Sale of tobacco to underage individuals is prohibited. However, there is no age restriction on smoking. Further, smoking in public places is prohibited under COTPA 2003. However, probably due to the displacement effect of this legislation on smoke-free public places, smokers choose smoking inside their houses, leading to a high prevalence of indoor SHS in India. A similar displacement effect of smoke-free legislation leading to increased indoor smoking and SHS was documented in Macao, China (30). To address these issues, India should fully comply with the FCTC requirements for a smoke-free environment in all indoor places. Additionally, other measures, such as increasing access to tobacco cessation services, more visible and aggressive anti-tobacco campaigns (73), and interventions promoting smoke-free homes, can play a pivotal role in reducing indoor SHS (11,74,75). The interventions of smoke-free homes, including providing face-to-face counselling and information to smokers, have shown a significant positive impact on reducing indoor smoking in other settings (76–81). Similar interventions can help reduce indoor SHS and overall tobacco use in India.

## Conclusion

The study concludes that the prevalence of indoor SHS exposure is alarmingly high among Indian children and non-smoker adults. Accordingly, every second child is exposed to indoor SHS. While every four out of ten non-smoking Indian adults are exposed to indoor SHS. Indoor SHS is prevalent in north-eastern, central, and northern states. Every six out of ten non-smoking household members from the NER are exposed to indoor SHS. While every second non-smoker individual, either from the north or central region of India, is exposed to indoor SHS. This suggests a need for priority attention for implementing tobacco control policies in these regions.

The predictability of indoor SHS exposure with respect to socio-demographic characteristics provided a better understanding of the prevalent yet least-studied issue of indoor SHS and has been a crucial step in identifying the at-risk population. Specifically, the younger age group (under 18), females, uneducated individuals, rural residents, occupants of *kuccha* houses, and poor households (poorest wealth index) are prone to indoor SHS exposure. Additionally, the susceptibility to indoor SHS exposure increases with an increase in the number of family members. There exists a hazardous intersection of age (childhood) and gender (female) specific susceptibility that is further intensified by the poor housing conditions with limited space (*Kaccha* houses), overcrowding, and poverty (poorest wealth index), ultimately leading to significantly high risk among the non-smoker household population with these attributes. This underscores the importance of community-based interventions creating awareness about tobacco use harms among the general public and providing behavior change communication (BCC) for smokers. Additionally, components of health education covering the effects of indoor tobacco SHS exposure should be included in the school curriculum. This will have a twin impact: the first, easy dissemination of health information through the channels of students to their parents and other adults in their household. Second, it can minimize the smoking susceptibility among school-going children. As schoolchildren aware of the health hazards of smoking won’t imitate smoking adults in their home and initiate smoking.

The study has few limitations attributed to study design and use of DHS data. First, there is the possibility of reporting bias arising from the self-reported data. Second, compared to longitudinal assessments, cross-sectional study design is not enough to establish strong causal relationships between indoor SHS and other socio-demographic factors specified in the study.

Third, the goodness-of-fit of the model was assessed using the Hosmer-Lemeshow test. The significant results (*p* < 0.05) showed that the model doesn’t fit the data well, suggesting the possibility of unexplored interaction effects between independent variables. However, in the absence of any national-level longitudinal study, the present study should be considered a notable attempt to present a brief overview of the issue.

Fourth, the data for the NFHS-5/DHS 2021, was collected during the COVID-19 pandemic period (Phase I: June 2019 to January 2020 and Phase II: January to April 2021) (42,82). Particularly the second phase of the survey was delayed during this period (83). Several studies have reported the impact of COVID-19 lockdown on overall human life and indicators captured by large scale surveys such as NFHS-5 (82–85). This is the major limitation of the current study as it can’t assess the actual impact of lockdown on indoor SHS exposure. It is acknowledged that due to COVID-19 lockdown, individuals mostly stayed at home and had prolonged exposure to indoor SHS. Possibly due to lockdown period, large number of household members, might have exposed to indoor SHS, contributing to higher prevalence of indoor SHS exposure than non-COVID period.

An integrated approach covering the above-discussed aspects of detailed assessment of the issue (covering intensity and longitudinal assessments), policy reforms restricting tobacco use in residential areas, and BCC-based targeted interventions focusing on smokers are crucial for smoke-free homes.

## Acknowledgments

The authors are grateful to the USAID, ICF, and Demographic and Health Surveys (DHS) Program, Rockville, MD 20850 USA, for authorizing the use of the India DHS 2021/NFHS-5 dataset for this study.

## Author Contributions

VT conceptualized and designed the entire study. VT was associated with the responsibilities of approaching data sources, obtaining authorization from DHS, data cleaning, data analysis, and presenting findings. VT also drafted the entire final draft of the manuscript, revised it, and finalized it for submission. KD applied formatting and page setup for the final draft and is the corresponding author of this paper. PG prepared a GIS map and assisted with the data analysis process. All authors approved the final version for submission.

## Data availability statement

The data that support the findings of this study can be requested from the DHS program at https://dhsprogram.com/Data/terms-of-use.cfm

## Biographical Notes

**Dr. Vishal Tikhute**

Dr. Vishal is Managing Director (MD) of Pragati Creations. Additionally, he oversees the R&D wing of the organization. For more than 11 years, Dr. Vishal has shared his expertise with researchers from prestigious international institutes (including Friedensau Adventist University, Germany, Linpico SARL - a French consultancy) from Africa and Southeast Asia. He is a public health professional with expertise in statistical analysis, program management, and scientific writing. Through his extensive international publications, he always advocates for quality healthcare delivery and human development for all sections of society.

**Ms. Komal Devkate**

Ms. Komal is a director at Pragati Creations; she looks after the community development and communications wing of the institute. She is visiting faculty at multiple universities and also associated with TISS, Mumbai, as a PhD scholar. Her research and project management experience exceeds 10 years of impressive work at various renowned institutes across India. Her research areas involved health equity, promoting women’s health rights, and improving healthcare access for rural, marginalized tribal communities, specifically adolescent girls and women from NTDNT communities.

**Mr. Prashant Gawai**

Mr. Prashant secured his master’s in Cities and Governance from the prestigious TISS, Hyderabad. Currently, he is a research consultant at Pragati Creations. He is also associated with multiple national and international institutes as a policy analyst. He contributes his expertise in health system strengthening by analyzing policies for gaps and exploring potential for reforms. He has command over social research, project management, and advanced data analytics tools. His areas of interest include sustainable human development through urban planning, housing, occupational safety, and mitigating environmental degradation.

